# Analysis of HER2-low breast cancer in Aotearoa New Zealand: a nationwide retrospective cohort study

**DOI:** 10.1101/2024.07.10.24310238

**Authors:** Annette Lasham, Reenadevi Ramsaroop, Abbey Wrigley, Nicholas Knowlton

**Affiliations:** Department of Molecular Medicine and Pathology, University of Auckland; Waitematā Hospital, Health NZ; Canopy Cancer Care; Department of Obstetrics and Gynaecology and Reproductive Sciences, University of Auckland; School of Mathematical and Computational Sciences, Massey University, Albany

## Abstract

**Aim:** To perform the first national analysis of demographic and clinicopathological features associated with the HER2 positive, HER2-low and HER2-zero invasive breast cancers in New Zealand. The study will inform the proportion of women who benefit from new HER2-targeted antibody drug conjugate (ADC) therapies.

**Methods:** Utilising data from Te Rēhita Mate Ūtaetae (Breast Cancer Foundation NZ National Register), the study analysed data from women diagnosed with invasive breast cancer over a 21-year period. The HER2 status of tumours was classified into three categories – HER2-zero, -low, - positive.

**Results:** From 2009-2021, 94% of women underwent HER2 testing, with 14% diagnosed with HER2-positive breast cancer. For advanced-stage disease, 38% formerly classified as HER2-negative were reclassified as HER2-low. Including HER2-positive breast cancers, this indicates 60% of women with advanced breast cancer would be eligible for the new HER2-directed ADCs (approximately 120 women per year). In future, these therapies may provide a targeted option for 40% of women with early-stage triple negative breast cancer now classified as HER2-low.

**Conclusion:** The findings suggest a significant proportion of women with invasive breast cancer in New Zealand could benefit from new HER2-targeted treatments. There is a need to standardise HER2 testing to enhance personalised treatment and improve outcomes.

## Introduction

Aotearoa New Zealand (NZ) has some of the highest rates of breast cancer in in the world. ^1^ Annually, NZ records approximately 3,500 new cases of breast cancer, with lifetime diagnosis of one in nine women.^2^ The mortality rates from breast cancer is around 650 each year.^2^ Notably, significant ethnic disparities in breast cancer incidence and mortality rates have been documented, particularly affecting wāhine Māori and Pacific women.^3–6^ Furthermore, compared to Australia, NZ experiences a 16% higher mortality rate from breast cancer, a discrepancy partly attributed to limited access to publicly-funded cancer medications in NZ.^7–9^ This is relevant as new targeted treatment options are readily available for breast cancer patients internationally.

A good example are drugs targeting tumours over-expressing the ERBB2/HER2 protein (henceforth called HER2). These HER2 positive tumours, represent 15-24% of breast cancers. 6 In NZ, trastuzumab was publicly funded for women with HER2 positive advanced breast cancer from July 2005, and subsequently for early breast cancer, initially for 9 weeks from July 2007 and later extended to 12 months from December 2008.^10^ Treatment is dependent on companion testing for overexpression of HER2 protein via immunohistochemistry (IHC) and *ERBB2* (HER2) gene amplification via *in situ* hybridisation (ISH), and has shown to produce benefit for patients considered HER2-positive as defined by IHC 3+ staining, or 2+ staining and ISH positive.

Recent clinical trials have demonstrated good outcomes in patients with lower levels of HER2 protein expression and/or without gene amplification. These tumours have previously been classified as “HER2 negative” but now are amenable for treatment with HER2-antibody drug conjugate (ADC), such as trastuzumab deruxtecan (T-DXd).^11^ These tumours are now categorised as HER2-low, distinct from those which have no HER2 expression (now called HER2-zero). The efficacy of this approach was demonstrated in DESTINY-Breast04 for patients with unresectable or metastatic HER2-low breast cancer, which showed significant improvement in progression-free survival and overall survival with T-DXd compared to standard chemotherapy.^11^ This now provides targeted treatment options for patients with both the newly-identified HER 2-low and HER2-positive tumours.

The aim of this paper is to perform the first national analysis of demographic and clinicopathological features associated with HER2-positive, HER2-low, and HER2-zero invasive breast cancers across New Zealand, to estimate the proportion of women who stand to benefit from new targeted therapies for HER2-low and HER2-positive tumours.

## Methods

### Data Source

Data extracted from Te Rēhita Mate Ūtaetae (Breast Cancer Foundation NZ National Register) included women diagnosed with breast cancer in Aotearoa New Zealand (NZ) between 2000– 2021. Te Rēhita is an opt-out registry that contains prospectively collected information on breast cancer diagnosed in Auckland and Waikato regions from 2000, in Christchurch region from 2009, the Wellington region from 2010 and nationally from 2020. The opt-out rate of 0.1% since 2012.^6^ Ethnicity information in Te Rēhita is populated from the Ministry of Health data, and this study followed the assignment of persons to single ethnicity groups as described in.^6^

### Variables

Only women with invasive breast cancer (AJCC7 stages 1-4) were eligible for this analysis (Figure 1). Women with invasive tumours that had missing hormone receptor data, or no HER2 data, or that were HER2 FISH negative with no HER2 immunohistochemistry (IHC) data were omitted from subsequent analyses of HER2 status (19%). Hormone receptor status of tumours was defined as follows-hormone receptor positive (HR+) was oestrogen receptor (ER) and/or progesterone receptor (PR) positive and hormone receptor negative (HR-) as ER and PR negative (following 1% cut off guidelines for ER/PR). HER2 status was defined as HER2 positive if the tumour was either FISH positive or IHC staining of 3+. HER2 negative tumours were defined as either FISH negative and/or IHC staining of 0, 1+ or 2+. HER2 low tumours were defined as FISH negative and/or IHC staining of 1+ or 2+. HER2-zero (HER2-0) tumours were defined as IHC staining of 0.

**Figure 1:**
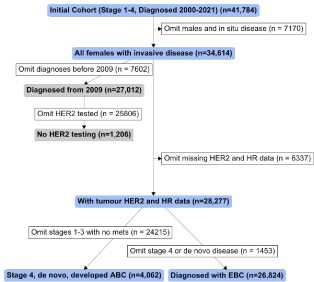
Flowchart showing inclusion (blue/grey boxes) and exclusion (white boxes) criteria for this study. Men diagnosed with breast cancer (n=287) and women diagnosed with *in situ* disease (stage 0, n= 6,528) were omitted at the outset. Only for the purpose of investigating which women did and did not receive HER2 tumour testing over time, distinct cohorts were defined comprising women with invasive breast cancer with or without HER2 tumour data (grey boxes). For the main analyses (shown with blue boxes), all women with invasive disease, with tumour hormone receptor (HR) and HER2 data (either HER2 positive by FISH, or with HER2 immunohistochemistry staining data) were included and then further analysed in cohorts of advanced breast cancer (ABC; diagnosed with Stage 4 disease, *de novo* disease or developed advanced breast cancer) and early breast cancer (EBC; diagnosed with Stages 1-3 disease). Note that 2,625 women diagnosed with EBC developed advanced breast cancer, so these women are included in both ABC and EBC cohorts.

### Statistical analyses

Data was summarised in counts (N) and percentages for categorical data. Differences of proportions between two or more subgroups were tested with Chi-Sq tests. A nominal alpha of 0.01 is considered statistically significant. All analyses were performed in R 4.3.1. Comparative analysis between advanced and early breast cancer cohorts were omitted owing to the lack of mutual exclusivity between these groups.

### Ethics

Te Rēhita Mate Ūtaetae operates under the NZ Health and Disability Ethics Committee approval (16/NTA/139/AM03), privacy and health legislation and Treaty of Waitangi principles.^6^ This specific study was approved by the Auckland Health Research Ethics Committee (AH2800).

## Results

### How many women had HER2 testing and who gets tested?

The HER2 test is a companion diagnostic test and is required to decide on treatment regimes. The proportion of women who underwent HER2 tumour testing, via either immunohistochemistry or ISH, starting from the year 2009 was analysed. This showed that approximately 94-96% of women had their breast tumours tested for HER2 (Table 1A). Of those tested, approximately 13-15% of women were diagnosed with HER2 positive breast cancer. Most regions performed 60-70% of HER2 testing on the excised lesion, except Midland, where nearly 90% of HER2 testing was performed on the core biopsy sample (Table 1B). Analysis by age at diagnosis showed that 2.3% of women under 70 years did not receive HER2 testing, but this was considerably higher at 13.1% for women diagnosed from 70 years and over (Table 1C). 23.1% of women under 45 years had HER2 positive breast tumours, in contrast to 13.3% of women diagnosed between 45-69 years. Approximately 6% of European women did not have HER2 testing, almost double that of wāhine Māori (Table 1D). As previously reported, ^6, 12^ a significantly higher proportion of Pacific women had HER2 positive breast cancers (21.5%) than European women (12%). Analysis by region of domicile at diagnosis revealed that 3-4% of women were not tested for HER2 status across all regions, with the exception of the Southern region, where over 10% of women’s tumours were untested for HER2 status (Table 1E). The proportion of women with HER2 positive breast tumours was higher in Northern and Southern regions.

**Table 1A:** Numbers and proportions of women who had tumour HER2 testing and testing results by their year of diagnosis.

| Year of Diagnosis | 2009-2011<br>(N=4,443) | 2012-2014<br>(N=5,538) | 2015-2017<br>(N=5,917) | 2018-2020<br>(N=7,646) | 2021<br>(N=3,468) |
| --- | --- | --- | --- | --- | --- |
| HER2 Negative | 3,586 (80.7%) | 4,473 (80.8%) | 4,771 (80.6%) | 6,382 (83.5%) | 2,865 (82.6%) |
| HER2 Positive | 606 (13.6%) | 743 (13.4%) | 873 (14.8%) | 950 (12.4%) | 431 (12.4%) |
| Not Tested | 251 (5.6%) | 322 (5.8%) | 273 (4.6%) | 314 (4.1%) | 172 (5.0%) |

**Table 1B:**
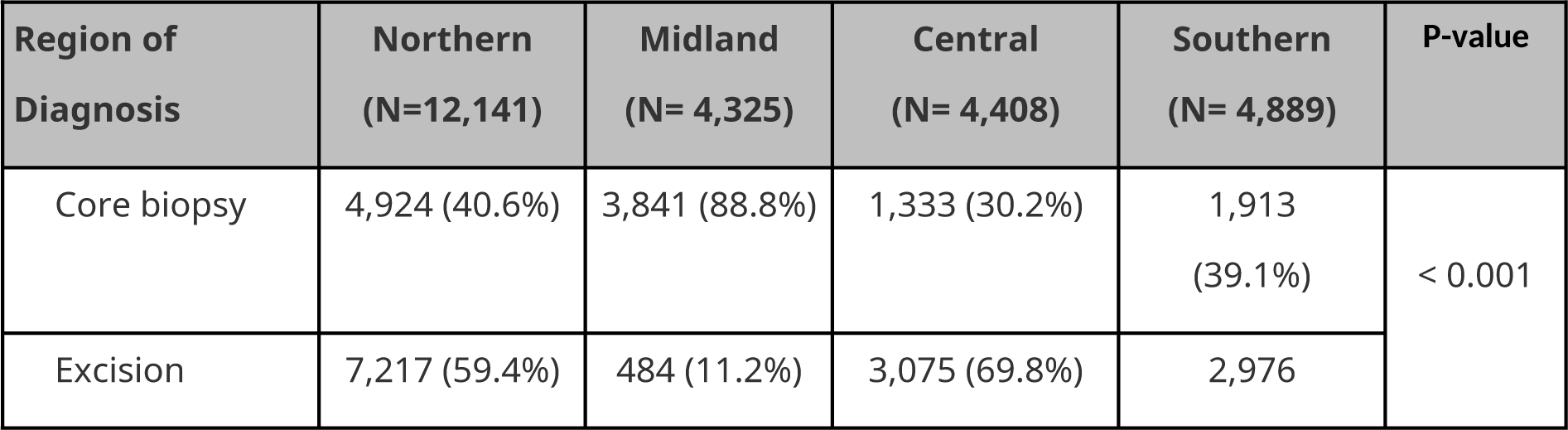
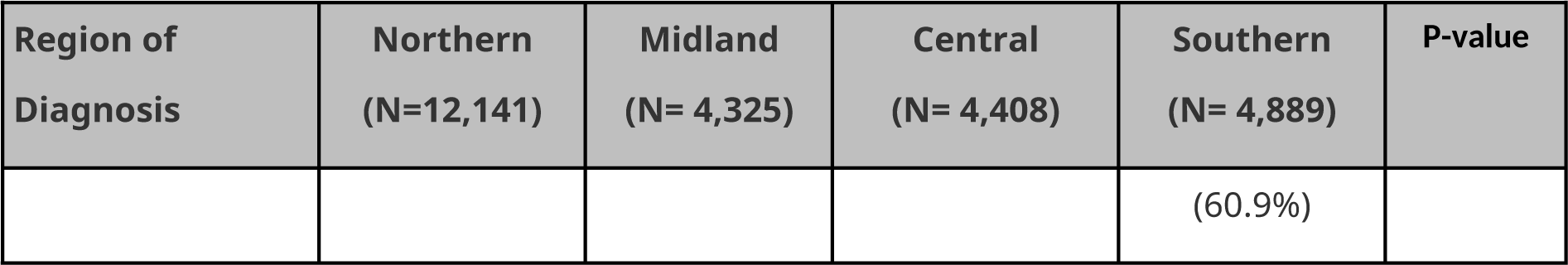
Sample used for HER2 testing by region of diagnosis (since 2009)

**Table 1C:** Numbers and proportions of women (diagnosed from 2009) who had tumour HER2 testing by their age at diagnosis.

| Age at Diagnosis | ≤ 44 years<br>(N = 3,122) | 45 - 69 years<br>(N=17,383) | ≥ 70 years<br>(N= 6,507) | P-value |
| --- | --- | --- | --- | --- |
| HER2 Negative | 2,332<br>(74.7%) | 14,666<br>(84.4%) | 5,079<br>(78.1%) | < 0.001 |
| HER2 Positive | 722 (23.1%) | 2,307 (13.3%) | 574 (8.8%) |  |
| Not Tested | 68 (2.2%) | 410 (2.4%) | 854 (13.1%) |  |

**Table 1D:** Numbers and proportions of women (diagnosed from 2009) who had tumour HER2 testing by their ethnicity.

| Ethnicity | Māori<br>(N= 3,131) | Pacific<br>(N= 1,765) | Asian<br>(N= 2,260) | Other<br>(N= 404) | European<br>(N=19,452) | P-value |
| --- | --- | --- | --- | --- | --- | --- |
| HER2 Negative | 2,560<br>(81.8%) | 1,313<br>(74.4%) | 1,828<br>(80.9%) | 329 (81.4%) | 16,047<br>(82.5%) | < 0.001 |
| HER2 Positive | 476 (15.2%) | 379 (21.5%) | 350 (15.5%) | 61 (15.1%) | 2,337<br>(12.0%) |  |
| Not Tested | 95 (3.0%) | 73 (4.1%) | 82 (3.6%) | 14 (3.5%) | 1,068 (5.5%) |  |

**Table 1E:** Numbers and proportions of women (diagnosed from 2009) who had tumour HER2 testing by the region they were diagnosed.

| Region of Diagnosis | Northern<br>(N=12,592) | Midland<br>(N= 4,405) | Central<br>(N= 4,549) | Southern<br>(N= 5,466) | P-value |
| --- | --- | --- | --- | --- | --- |
| HER2 Negative | 10,408 (82.7%) | 3,668 (83.3%) | 3,868 (85.0%) | 4,133<br>(75.6%) | 0.007 |
| HER2 Positive | 1,735 (13.8%) | 583 (13.2%) | 531 (11.7%) | 754 (13.8%) |  |
| Not Tested | 449 (3.6%) | 154 (3.5%) | 150 (3.3%) | 579 (10.6%) |  |

### Who was not tested?

Subsequent analysis was performed to understand which women did not have their tumours tested for HER2 status. Sixty seven percent of these women were over 69 years when diagnosed with breast cancer, and 81% were European women (Table 2). Regionally, the highest numbers of tumours that were not tested were in the Northern and Southern regions. Thirty-nine percent had stage 1 disease, while 27% had missing stage information but were documented as having invasive disease from a core biopsy sample (Table 2).

**Table 2:** Description of those women not receiving testing for tumour HER2 status, from 2009.

|  | Not<br>Tested<br>(N=1,206) |
| --- | --- |
| **Age at diagnosis** |  |
| ≤ 44 years | 57 (4.7%) |

|  | **Not**<br>**Tested**<br>**(N=1,206)** |
| --- | --- |
| 45 – 69 years | 336 (27.9%) |
| ≥ 70 years | 813 (67.4%) |
| **Ethnicity** |  |
| Māori | 74 (6.1%) |
| Pacific | 69 (5.7%) |
| Asian | 71 (5.9%) |
| Other | 14 (1.2%) |
| European | 978<br>(81.1%) |
| **Region of diagnosis** |  |
| Northern | 431<br>(35.7%) |
| Midland | 76 (6.3%) |
| Central | 136<br>(11.3%) |
| Southern | 563<br>(46.7%) |
| **TNM Stage** |  |
| 1 | 465<br>(38.6%) |
| 2 | 245<br>(20.3%) |
| 3 | 55 (4.6%) |
| 4 | 120 |
|  | (10.0%) |
| None<br>recorded | 321<br>(26.6%) |

### What is the proportion of HER2+, HER2-low and HER2-zero breast cancers in NZ?

The Register collects detailed data around the reporting of HER2 results and this enabled the HER2 negative tumours to be stratified into HER2-zero and HER2-low (acknowledging limitations).^13–15^ A further stratification via HR status divided the patient groups into HR positive and HR negative, which were then analysed based on whether the patients had advanced or early-stage breast cancer, allowing for a more detailed understanding of treatment needs across different stages of the disease.

#### Advanced Breast Cancer

Thirty one percent of women with HER2 negative advanced breast cancer could be re-categorised as HER2-low, 51% were HR+ and 39% triple negative (Figure 2).

**Figure 2.**
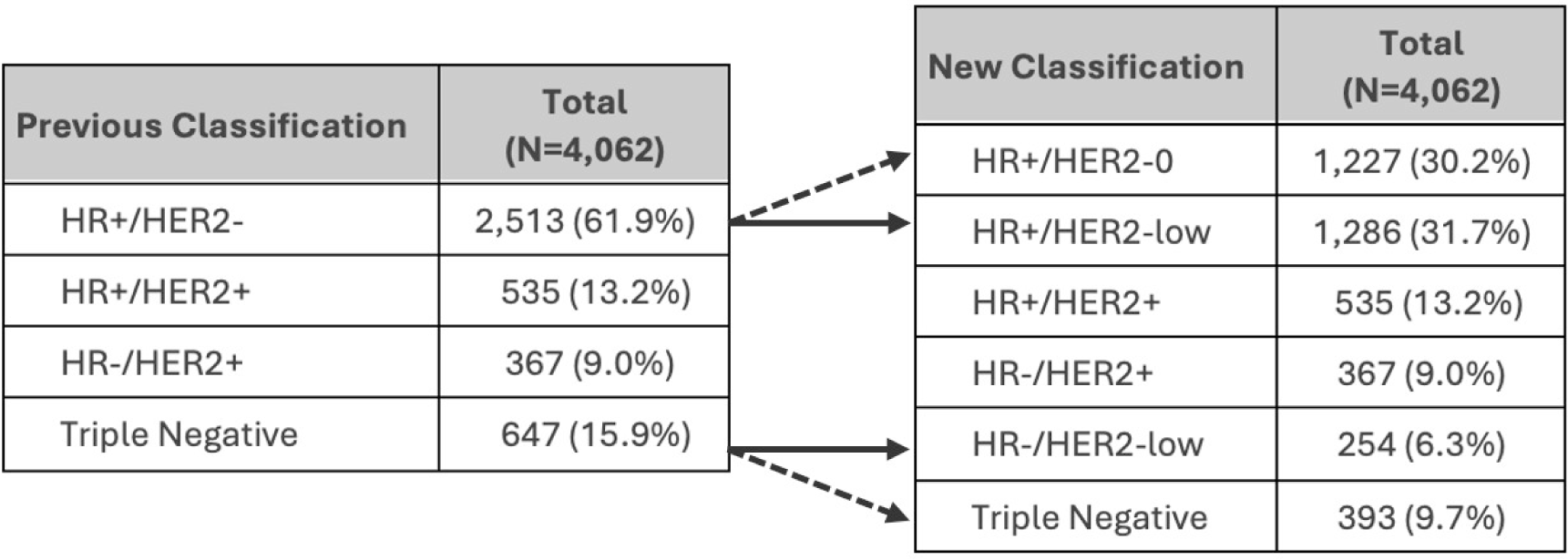
Analysis of women with advanced breast cancer by tumour receptor status, prior to and after recategorisation of HER2 negative tumours. HR = hormone receptor. Note: Arrows illustrate the recategorised patients according to the updated classification system. Triple Negative = HR-/HER2-0 in the right table.

Analysis of the six HER2 subclasses by demographic and pathological features for advanced disease revealed notable differences. Women diagnosed before the age of 45 had the highest proportions of HER2+ breast cancers, making this younger age group the most likely to benefit from HER-targeting ADC treatments (Table 3A). No statistical relationship was found between

HER2-low status and age group. By ethnicity, Pacific women displayed the highest proportion of HER2+ breast cancers (Table 3B). The incidence of HER2-low cancers was similar across all ethnic groups. While wāhine Māori and Pacific women would benefit disproportionately from access to HER2-targeting therapies, this advantage is primarily attributed to the higher prevalence of HER2-positive cases rather than an increased occurrence of HER2-low tumours. Analysis by tumour pathology revealed that HR-/HER2-low tumours were predominantly grade 3, while HR+/HER2-low were primarily grades 1 or 2 (Table 3C). Interestingly, the distribution of tumour sizes showed no significant variation across the subclasses.

**Table 3A:** Description of women with advanced breast cancer by HER2 subclasses by age at diagnosis.

| Age at Diagnosis | ≤ 44 years<br>(N=884) | 45-69 years<br>(N=2126) | ≥ 70 years<br>(N=1052) | P-value |
| --- | --- | --- | --- | --- |
| **Receptor Status** |  |  |  |  |
| HR+/HER2-0 | 231 (26.1%) | 634 (29.8%) | 362 (34.4%) | ns |
| HR+/HER2-low | 266 (30.1%) | 671 (31.6%) | 349 (33.2%) |  |
| HR+/HER2+ | 148 (16.7%) | 275 (12.9%) | 112 (10.6%) |  |
| HR-/HER2+ | 104 (11.8%) | 201 (9.5%) | 62 (5.9%) |  |
| HR-/HER2-low | 49 (5.5%) | 138 (6.5%) | 67 (6.4%) |  |
| Triple Negative | 86 (9.7%) | 207 (9.7%) | 100 (9.5%) |  |
| **Total that would benefit from HER-targeting ADC treatments** | 567 (64%) | 1285 (60%) | 590 (56%) |  |

**Table 3B:** Description of women with advanced breast cancer by HER2 subclass by ethnicity.

| Ethnicity | Māori<br>(N=473) | Pacific<br>(N=420) | Asian<br>(N=286) | European<br>(N=2759) | P-value |
| --- | --- | --- | --- | --- | --- |
| **Receptor Status** |  |  |  |  |  |
| HR+/HER2-0 | 133 (28.1%) | 119 (28.3%) | 80 (28.0%) | 866 (31.4%) | 0.010 |
| HR+/HER2-low | 155 (32.8%) | 138 (32.9%) | 87 (30.4%) | 866 (31.4%) |  |
| HR+/HER2+ | 74 (15.6%) | 80 (19.0%) | 39 (13.6%) | 327 (11.9%) |  |
| HR-/HER2+ | 47 (9.9%) | 54 (12.9%) | 34 (11.9%) | 212 (7.7%) |  |
| HR-/HER2-low | 28 (5.9%) | 13 (3.1%) | 15 (5.2%) | 190 (6.9%) |  |
| Triple Negative | 36 (7.6%) | 16 (3.8%) | 31 (10.8%) | 298 (10.8%) |  |
| **Total that would benefit from HER-targeting ADC treatments** | 304 (64%) | 285 (67%) | 176 (62%) | 1595 (58%) |  |

124 women with "other ethnicity" omitted from this table due to low numbers across the subclasses

**Table 3C:** Description of advanced tumours by HER2 subclasses – Grade and tumour size.

| Tumour<br>Receptor<br>Subtype | HR+/<br>HER2-0<br>(N=1227) | HR+/<br>HER2-low<br>(N=1286) | HR+/<br>HER2+<br>(N=535) | HR-/<br>HER2+<br>(N=367) | HR-/<br>HER2-low<br>(N=254) | Triple<br>Negative<br>(N=393) | P-value |
| --- | --- | --- | --- | --- | --- | --- | --- |
| Tumour grade |  |  |  |  |  |  |  |
| 1 or 2 | 592<br>(48.2%) | 615<br>(47.8%) | 129<br>(24.1%) | 47<br>(12.8%) | 34<br>(13.4%) | 58<br>(14.7%) | <0.001 |
| 3 | 335<br>(27.3%) | 401<br>(31.2%) | 250<br>(46.7%) | 217<br>(59.1%) | 187<br>(73.6%) | 287<br>(73.0%) |  |
| None<br>recorded | 300<br>(24.5%) | 270<br>(21.0%) | 156<br>(29.1%) | 103<br>(28.1%) | 33<br>(13.0%) | 48<br>(12.2%) |  |
| Tumour size |  |  |  |  |  |  |  |
| ≤ 20 mm | 192<br>(15.6%) | 216<br>(16.8%) | 92<br>(17.2%) | 74<br>(20.2%) | 49<br>(19.3%) | 87<br>(22.1%) | ns |
| 21 – 50<br>mm | 565<br>(46.0%) | 571<br>(44.4%) | 219<br>(40.9%) | 149<br>(40.6%) | 131<br>(51.6%) | 204<br>(51.9%) |  |
| > 50 mm | 172<br>(14.0%) | 214<br>(16.6%) | 73<br>(13.6%) | 51<br>(13.9%) | 41<br>(16.1%) | 59<br>(15.0%) |  |
| None | 298 | 285 | 151 | 93 | 33 | 43 |  |

| Tumour Receptor Subtype | HR+/HER2-0 (N=1227) | HR+/HER2-low (N=1286) | HR+/HER2+ (N=535) | HR-/HER2+ (N=367) | HR-/HER2-low (N=254) | Triple Negative (N=393) | P-value |
| --- | --- | --- | --- | --- | --- | --- | --- |
| recorded | (24.3%) | (22.2%) | (28.2%) | (25.3%) | (13.0%) | (10.9%) |  |

Tumour grade and size determined from histopathology data.
Tumour grades 1 and 2 were combined because of low numbers for some subclasses. "ns" = not significant

#### Early breast cancer

Although clinical trials have not yet been conducted for early-stage breast cancer, we examined what proportion of women had HER2-low tumours. Our analysis showed that 43% of women with HER2-negative early breast cancer could be reclassified as having HER2-low disease. Specifically, this includes 51% of those previously classified as HR+/HER2-negative and 40% of those previously identified as triple-negative early breast cancers (Figure 3).

**Figure 3.**
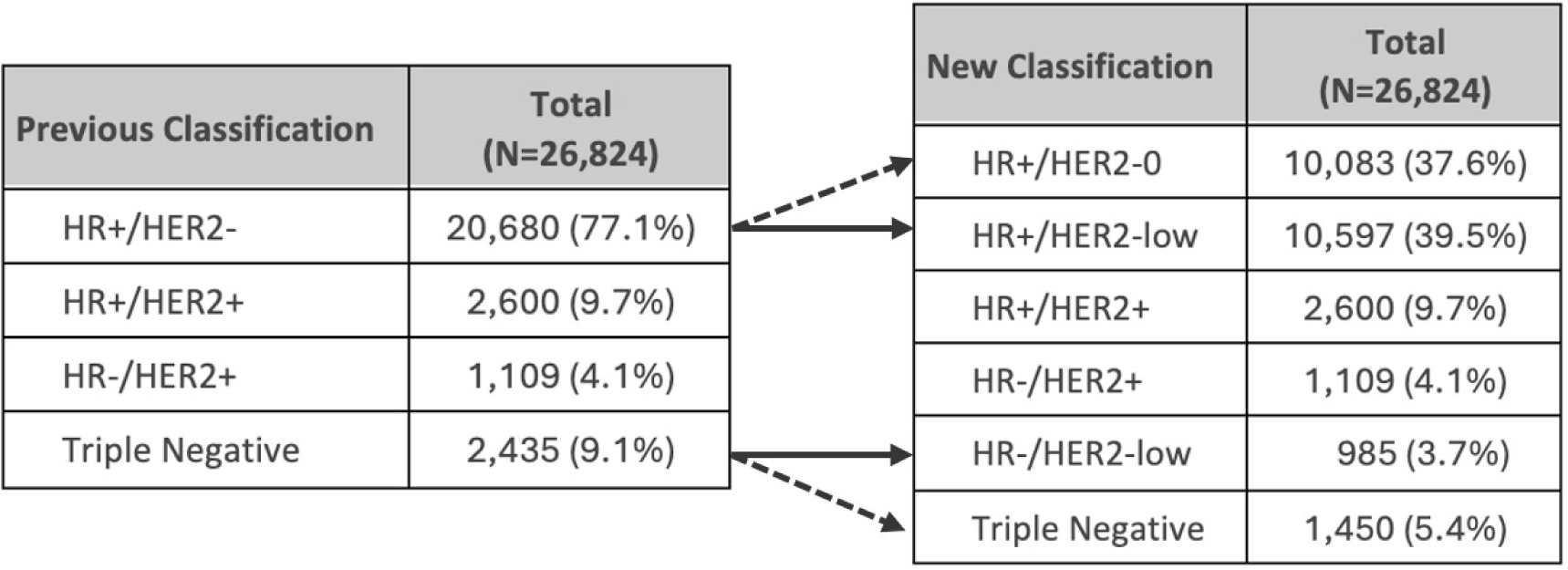
Analysis of women with early breast cancer by tumour receptor status, prior to and after reclassification of HER2 negative tumours. HR = hormone receptor. Note: Arrows illustrate the reclassification of patients according to the updated classification system. Triple Negative = HR-/HER2-0 in the right table.

Analysis of the various receptor subclasses by women’s demographics showed that those diagnosed before 45 years of age had the highest proportions of tumours with poor prognostic features (HR+/HER2+ and HR-tumours 16% and 20% respectively), compared to women diagnosed from age 45 years (who had 9% of HR+/HER2+ and 12% HR-tumours) (Table 4A). By ethnicity, 22% of Pacific women were diagnosed with HER2-positive breast cancers, compared to 13-16% among women of other ethnicities (Table 4B). Apart from this, the distribution of each subclass was similar across ethnic groups. Irrespective of the age at diagnosis or ethnicity, 40% of women with tumours previously classified as triple negative would now be classified as HR-/HER2-low. Analysis of the subclasses by clinicopathological features revealed that HR+/HER2-low and HR+/HER2-zero breast cancers had similar characteristics: 59-60% were stage 1, 63% had N0 nodal status, 50-51% were grade 2 tumours, and 55% of tumours measured 20mm or smaller (Table 4C). Similarly, HR-/HER2-low and triple-negative breast cancers also showed comparable features with 48% being stage 1, 43-44% stage 2, 77-78% grade 3 tumours, and 48-49% measuring 21–50 mm in size, although nodal status was more variable (Table 4C).

**Table 4A:** Description of women diagnosed with early breast cancer by HER2 subclasses by age at diagnosis.

| Age at Diagnosis | ≤ 44 years<br>(N=3512) | 45-69 years<br>(N=17889) | ≥ 70 years<br>(N=5423) | P-value |
| --- | --- | --- | --- | --- |
| **Receptor Status** |  |  |  |  |
| HR+/HER2-0 | 1050 (29.9%) | 6792 (38.0%) | 2241 (41.3%) | <0.001 |
| HR+/HER2-low | 1219 (34.7%) | 7225 (40.4%) | 2153 (39.7%) |  |
| HR+/HER2+ | 553 (15.7%) | 1664 (9.3%) | 383 (7.1%) |  |
| HR-/HER2+ | 218 (6.2%) | 747 (4.2%) | 144 (2.7%) |  |
| HR-/HER2-low | 190 (5.4%) | 595 (3.3%) | 200 (3.7%) |  |
| Triple Negative | 282 (8.0%) | 866 (4.8%) | 302 (5.6%) |  |

**Table 4B:** Description of women diagnosed with early breast cancer by HER2 subclasses by ethnicity.

| Ethnicity | Māori<br>(N=2882) | Pacific<br>(N=1743) | Asian<br>(N=2321) | Other<br>Ethnicity<br>(N=435) | European<br>(N=19443) | P-value |
| --- | --- | --- | --- | --- | --- | --- |
| **Receptor Status** |  |  |  |  |  |  |
| HR+/HER2-0 | 1143<br>(39.7%) | 669 (38.4%) | 829 (35.7%) | 148 (34.0%) | 7294<br>(37.5%) | <0.001 |
| HR+/HER2-low | 1111<br>(38.5%) | 586 (33.6%) | 913 (39.3%) | 166 (38.2%) | 7821<br>(40.2%) |  |
| HR+/HER2+ | 297 (10.3%) | 239 (13.7%) | 262 (11.3%) | 47 (10.8%) | 1755 (9.0%) |  |
| HR-/HER2+ | 131 (4.5%) | 148 (8.5%) | 107 (4.6%) | 22 (5.1%) | 701 (3.6%) |  |
| HR-/HER2-low | 83 (2.9%) | 38 (2.2%) | 87 (3.7%) | 16 (3.7%) | 761 (3.9%) |  |
| Triple Negative | 117 (4.1%) | 63 (3.6%) | 123 (5.3%) | 36 (8.3%) | 1111 (5.7%) |  |

**Table 4C:**
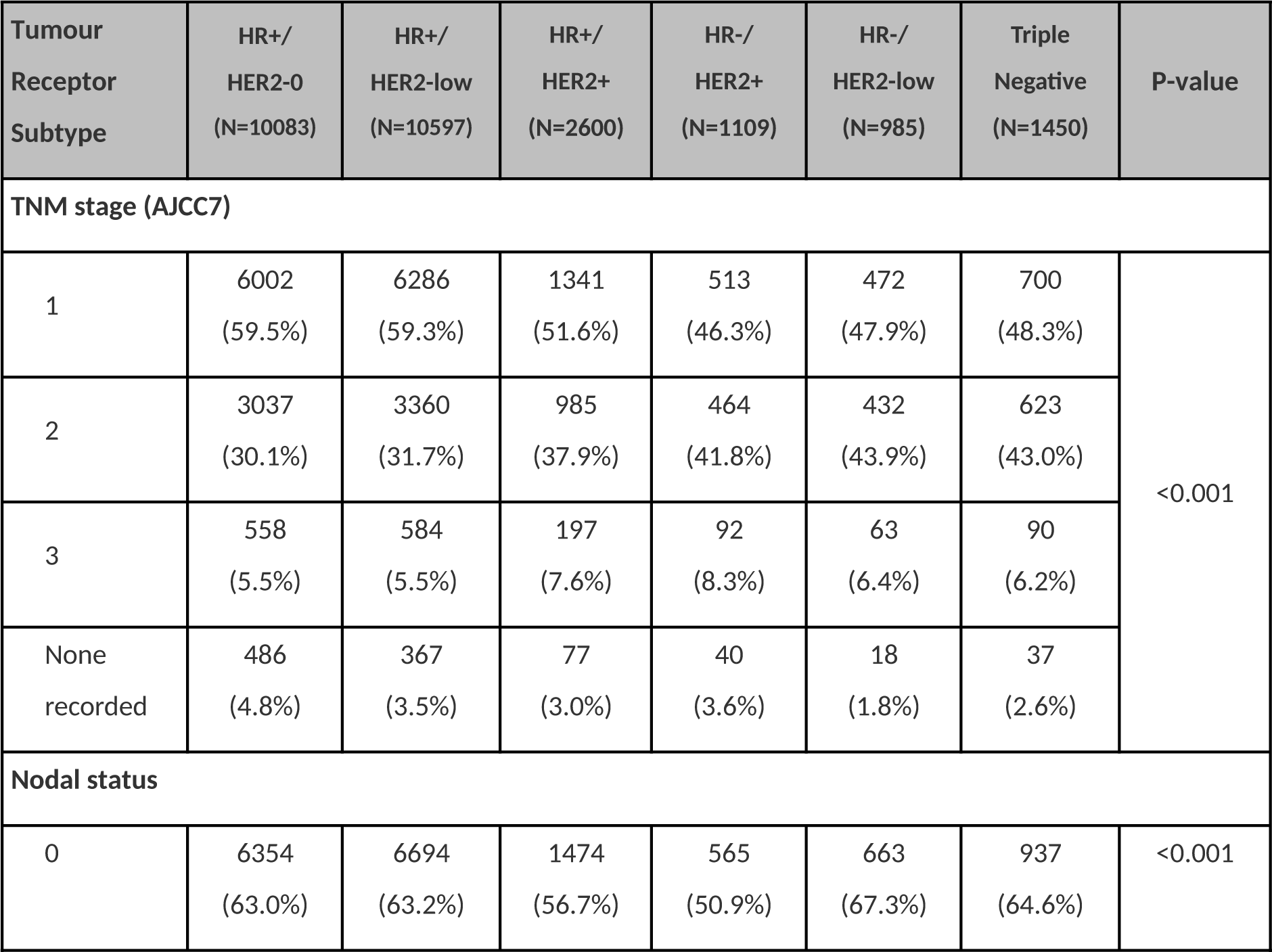
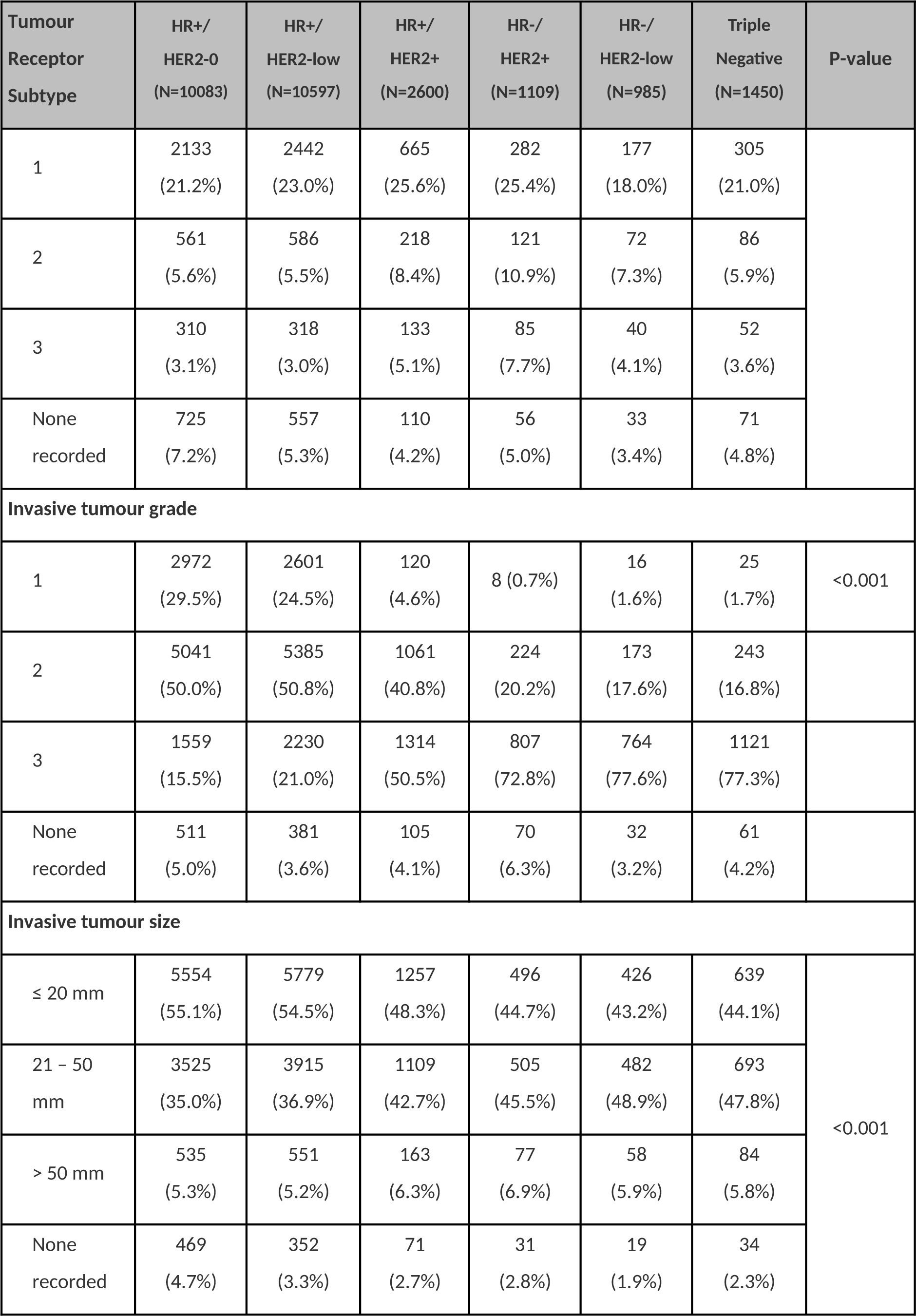
Description of clinicopathological features of early breast cancers by HER2 subclasses – stage, grade and size.

| Tumour<br>Receptor<br>Subtype | HR+/<br>HER2-0<br>(N=10083) | HR+/<br>HER2-low<br>(N=10597) | HR+/<br>HER2+<br>(N=2600) | HR-/<br>HER2+<br>(N=1109) | HR-/<br>HER2-low<br>(N=985) | Triple<br>Negative<br>(N=1450) | P-value |
| --- | --- | --- | --- | --- | --- | --- | --- |
| TNM stage (AJCC7) |  |  |  |  |  |  |  |
| 1 | 6002<br>(59.5%) | 6286<br>(59.3%) | 1341<br>(51.6%) | 513<br>(46.3%) | 472<br>(47.9%) | 700<br>(48.3%) | <0.001 |
| 2 | 3037<br>(30.1%) | 3360<br>(31.7%) | 985<br>(37.9%) | 464<br>(41.8%) | 432<br>(43.9%) | 623<br>(43.0%) |  |
| 3 | 558<br>(5.5%) | 584<br>(5.5%) | 197<br>(7.6%) | 92<br>(8.3%) | 63<br>(6.4%) | 90<br>(6.2%) |  |
| None<br>recorded | 486<br>(4.8%) | 367<br>(3.5%) | 77<br>(3.0%) | 40<br>(3.6%) | 18<br>(1.8%) | 37<br>(2.6%) |  |
| Nodal status |  |  |  |  |  |  |  |
| 0 | 6354<br>(63.0%) | 6694<br>(63.2%) | 1474<br>(56.7%) | 565<br>(50.9%) | 663<br>(67.3%) | 937<br>(64.6%) | <0.001 |
| 1 | 2133<br>(21.2%) | 2442<br>(23.0%) | 665<br>(25.6%) | 282<br>(25.4%) | 177<br>(18.0%) | 305<br>(21.0%) |  |
| 2 | 561<br>(5.6%) | 586<br>(5.5%) | 218<br>(8.4%) | 121<br>(10.9%) | 72<br>(7.3%) | 86<br>(5.9%) |  |
| 3 | 310<br>(3.1%) | 318<br>(3.0%) | 133<br>(5.1%) | 85<br>(7.7%) | 40<br>(4.1%) | 52<br>(3.6%) |  |
| None<br>recorded | 725<br>(7.2%) | 557<br>(5.3%) | 110<br>(4.2%) | 56<br>(5.0%) | 33<br>(3.4%) | 71<br>(4.8%) |  |
| Invasive tumour grade |  |  |  |  |  |  |  |
| 1 | 2972<br>(29.5%) | 2601<br>(24.5%) | 120<br>(4.6%) | 8 (0.7%) | 16<br>(1.6%) | 25<br>(1.7%) | <0.001 |
| 2 | 5041<br>(50.0%) | 5385<br>(50.8%) | 1061<br>(40.8%) | 224<br>(20.2%) | 173<br>(17.6%) | 243<br>(16.8%) |  |
| 3 | 1559<br>(15.5%) | 2230<br>(21.0%) | 1314<br>(50.5%) | 807<br>(72.8%) | 764<br>(77.6%) | 1121<br>(77.3%) |  |
| None<br>recorded | 511<br>(5.0%) | 381<br>(3.6%) | 105<br>(4.1%) | 70<br>(6.3%) | 32<br>(3.2%) | 61<br>(4.2%) |  |
| Invasive tumour size |  |  |  |  |  |  |  |
| ≤ 20 mm | 5554<br>(55.1%) | 5779<br>(54.5%) | 1257<br>(48.3%) | 496<br>(44.7%) | 426<br>(43.2%) | 639<br>(44.1%) | <0.001 |
| 21 – 50<br>mm | 3525<br>(35.0%) | 3915<br>(36.9%) | 1109<br>(42.7%) | 505<br>(45.5%) | 482<br>(48.9%) | 693<br>(47.8%) |  |
| > 50 mm | 535<br>(5.3%) | 551<br>(5.2%) | 163<br>(6.3%) | 77<br>(6.9%) | 58<br>(5.9%) | 84<br>(5.8%) |  |
| None<br>recorded | 469<br>(4.7%) | 352<br>(3.3%) | 71<br>(2.7%) | 31<br>(2.8%) | 19<br>(1.9%) | 34<br>(2.3%) |  |

## Discussion

The introduction of trastuzumab deruxtecan (T-DXd) marks a significant advancement in the personalised treatment landscape for women with breast cancer, particularly for those classified as having HER2-low tumours.^11^ This study provides the first analysis of HER2 testing patterns women diagnosed with breast cancer in Aotearoa New Zealand and the proportion of women who might benefit from these new drugs.

This analysis showed that from 2009, over 94% of women diagnosed with invasive breast cancer had HER2 testing, following international standards and establishing the critical role of HER2 testing in guiding therapies offered to NZ women. However, considerably lower testing rates were observed for women diagnosed after 70 years of age, of which 85% were European. These findings are consistent with a previous NZ study showing that chemotherapy was given to only 5% of women diagnosed with invasive breast cancer from age 70 years.^16^

The data also showed that women diagnosed with advanced breast cancers before the age of 45 exhibited the highest proportions of HER2+ tumours. Additionally, wāhine Māori and Pacific women with advanced disease displayed higher levels of HR+/HER2+. Notably, Pacific women recorded the highest proportion of HER2+ tumours across all disease stages, confirming previous findings.^6, 12^ For these groups, especially those with advanced breast cancer, access to new HER2-targeting ADCs could significantly enhance treatment efficacy and patient outcomes. Interestingly, our analysis revealed no significant differences in the age or ethnicity of patients with HER2-low tumours, suggesting that HER2-low status may manifest independently of these demographic factors.

The new stratification of breast cancers from HER2-negative to HER2-low or HER2-zero subclasses (noting that HR-/HER2-zero = triple negative) in our cohort has shown that HER2-low subclasses comprise over 50% of HR+/HER2-negative and ∼40% of triple negative breast cancers for both advanced and early breast cancers. Using this information, these new therapies could provide treatment options for 32% of advanced breast cancers previously classified as HER2-negative. In 2022, the FDA approved the use of T-DXd for unresectable or metastatic HER2-low breast cancer as second line therapy after chemotherapy, or if there was disease recurrence during or within six months of completing adjuvant chemotherapy.^17^ In addition to HER2-positive advanced breast cancer, this would be approximately 120 women per year based on 2020 and 2021 data that would be eligible in NZ.

At the time of writing, there are a number of clinical trials underway, evaluating T-DXd as a first line treatment, alone or in combination with pertuzumab, for HER2-positive advanced breast cancer (DESTINY-Breast07)^18^ and DESTINY-Breast06 which directly compares standard-of-care chemotherapy to T-DXd for HR+/HER2-low or HR+/HER2-ultralow (defined as IHC 0 with minimal (<1+) membrane staining) advanced breast cancers after one or more lines of endocrine therapy. To date, both of these trials have shown improved outcomes for patients given T-DXd.^19^

For future applications of HER2-ADCs for early-stage HER2-low breast cancer, in our opinion the most likely candidates are those with triple negative disease who are now defined as HR-/HER2-low. These women would then have a targeted therapy available, potentially transforming their treatment options. These drugs could serve as a targeted therapy for 40% of women formerly diagnosed with early triple negative breast cancer now recategorised as HER2-low, representing approximately 48 women per year in NZ based on 2020 and 2021 data.

Limitations: the main limitation is the classification of HER2-low and HER2-zero groups from the recorded HER2 immunohistochemistry data. As many studies by pathologists have emphasised, there is evidence of undercalling and overcalling of HER-2 staining level, in part because this level of detail has not previously required, this test was designed to produce a binary output-HER2-positive or HER2-negative.^13, 15^ In addition, with potential heterogeneity across a tumour sample, variable staining and reporting between labs, the stratification of our cohort into HER2-low and HER2-zero groups may not be robust.^13, 15^ These limitations apply going forward, hence why there are international efforts to generate standardised testing protocols to ensure reliable and actionable HER2 status determination.^14, 20^ Future advancements in diagnostic technologies aim to reduce these inconsistencies, providing a more reliable basis for treatment decisions.

The potential impact of these therapies for breast cancer in NZ is considerable, especially when considering the current limitations in drug funding and approval timelines by Pharmac.^7, 9^ An Australian study showed how access to systemic therapies saved the lives of women diagnosed with breast cancer,^21^ therefore ensuring access to these new therapies could help mitigate the observed excess mortality in breast cancer patients in NZ compared to Australia.

In conclusion, the potential to treat a wider spectrum of breast cancers with targeted therapies like T-DXd could transform the therapeutic landscape in New Zealand. Ensuring that these benefits reach all segments of the population will require concerted efforts in medical research, healthcare policy, and clinical practice. Our study sets the groundwork for these developments, advocating for a paradigm shift in how breast cancer is treated in the national context.

## Data Availability

All data produced in the present study are available upon reasonable request to Te Rēhita Mate Ūtaetae.

## Acknowledgements

We gratefully acknowledge Te Rēhita Mate Ūtaetae-Breast Cancer Foundation National Register, for providing the data used in this study. Our thanks also extend to all the individuals who consented to participate in the registry, contributing invaluable information towards breast cancer research and management. Special thanks to our colleagues who provided insight and expertise that greatly assisted the research.

